# Overdose-related communication in Connecticut: reported receipt, perceived utility, and responses among overdose prevention and response professionals

**DOI:** 10.64898/2026.10.01.26363949

**Authors:** A Ram, Frederick L. Altice

**Affiliations:** Division of Infectious Diseases, Department of Medicine, Yale School of Medicine, New Haven, Connecticut, USA; Program in Computational Biology and Biomedical Informatics, Yale University, New Haven, Connecticut, USA; Center for Interdisciplinary Research on AIDS, Yale School of Public Health, New Haven, Connecticut, USA; Department of Epidemiology of Microbial Diseases, Yale School of Public Health, New Haven, Connecticut, USA

**Keywords:** Overdose prevention, ODMAP, Connecticut, alerts, overdose

## Abstract

**Background:** Timely overdose response depends not only on surveillance systems that detect overdose clusters, but also on communication pathways that deliver actionable information to the professionals positioned to respond. Understanding which overdose-related communications are received, perceived as useful, and translated into action is essential for improving overdose prevention and response infrastructure.

**Objective:** We assessed which overdose-related communication channels professionals reported receiving, which they perceived as most helpful, how organizations reported responding to overdoses or overdose clusters, and whether these patterns differed by professional role, county, and funding source.

**Methods:** We conducted an exploratory descriptive survey in 2024 using a convenience sample of professionals across Connecticut whose jobs involved overdose prevention or response (N=149). Using this survey data, we summarized reported receipt and perceived utility of overdose-related communication channels, organizational response actions, and internal receipt of Connecticut Department of Public Health overdose alerts. Results were stratified descriptively by professional role, county, and funding source. We interpreted findings using a public health communication-to-action cascade focused on signal detection, communication receipt, internal dissemination, perceived utility, and response action.

**Results:** Reported communication receipt and perceived utility differed most clearly by professional role. Peer information sharing was the most commonly reported communication source and was frequently perceived as helpful, but respondents who reported receiving formal channels such as state alerts, local alerts, or platform-based information often identified those channels as most helpful. Internal receipt of Connecticut Department of Public Health overdose alerts also varied by role. SSPs/substance use treatment programs and first responders more often reported outreach to people who use drugs, whereas government and health district respondents reported a more even mix of public communication and outreach activities.

**Conclusions:** Overdose-related communication in Connecticut appears role-dependent and unevenly distributed. Informal peer networks are widely used and perceived as helpful, while formal alert systems may require improvements to reach and actionability. Practical improvements include opt-in communication pathways tailored to professional roles, clearer access points for state and local alerts, internal dissemination protocols that specify who should receive which alerts, and alert-to-action playbooks that connect specific alert types to recommended response steps.

## Introduction

Overdose prevention and response depend on timely situational awareness and effective communication across public health, harm reduction, treatment, emergency response, and community stakeholders. Jurisdictions use a mix of mechanisms to disseminate overdose-related information, including formal alerts, dashboards, listservs, and informal peer-to-peer networks. Effective overdose responses require rapid action, and overdose prevention efforts require planning and coordination.^1–3^ The more efficient and well-utilized a communication system is, the more able it is to help professionals respond to and prevent overdoses.^4^

One widely used surveillance tool is the Overdose Detection Mapping Application Program [ODMAP], which supports near-real-time monitoring of suspected overdose events and can generate “spike alerts” when reported overdoses meet or exceed expected thresholds.^5,6^ The public health value of such alerts depends on the downstream communication pathway: who receives them, whether recipients perceive them as useful, whether alerts are disseminated within organizations, and whether they are linked to feasible overdose responses.

In Connecticut, the Department of Public Health [CT DPH] uses ODMAP spike alerts to notify professionals of emerging crises and prompt vigilance and rapid action.^7^ During a 2019 cluster of overdoses in Hartford, this system allowed the state to quickly identify the location of overdoses, which then helped them facilitate a coordinated public health response involving multiple agencies that identified the substances involved and took steps to curb subsequent overdoses.^8^ Recent after-action reflections from state partners describe similar rapid deployments during 2025 surges.^9^

The broader communication ecosystem, however, extends beyond ODMAP and includes multiple channels that vary in accessibility, timeliness, and perceived utility across stakeholder groups. Understanding who receives which communications, through what channels, and what actions follow is essential for improving the effectiveness and impact of overdose response efforts. Additionally, there is little comprehensive documentation of how overdose responses as motivated by these communication efforts may vary by location and profession. To organize this communication-to-action problem, we conceptualize overdose response communication as a cascade linking overdose event detection, alert generation, organizational receipt, internal dissemination, perceived utility/actionability, and response action **[Figure 1].**

**Figure 1:**
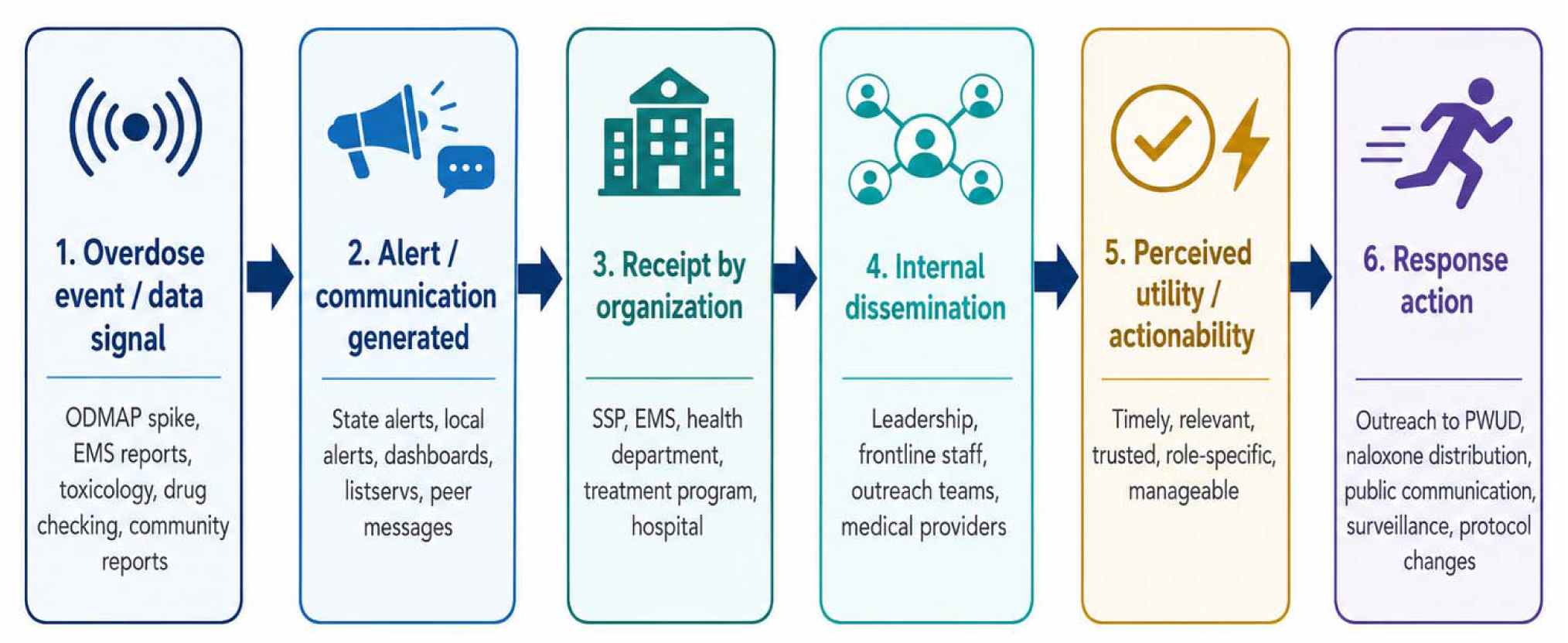
Conceptual communication-to-action cascade for overdose response

In this study, we surveyed 149 professionals involved in overdose prevention/response across Connecticut to characterize who receives different forms of overdose communication and by which channels, which channels they find to be most helpful, and what actions are taken during an overdose crisis. We characterized overdose-related communication as a public health communication-to-action pathway: receipt of information, perceived utility of communication channels, internal dissemination of state alerts, and reported responses. We also explored whether these descriptive patterns varied by professional role, county, and funding source. Finally, we examined gaps between communication receipt and perceived utility to generate practical recommendations for strengthening overdose-related communication systems, using constructs from Proctor’s Implementation Outcomes as an evaluative framework.^10^

## Methods

### Ethics statement

This study was approved by the Yale Human Research Protection Program [HRPP]. Yale HRPP deemed that this study met the federal and university criteria for exemption [approved protocol ID: 2000034327]. We received implied participant consent via survey completion, as participation was voluntary. To reduce barriers to participation or potential concerns about professional sensitivity or stigma, the survey was anonymous, did not collect IP addresses and did not require contact information. This study is STROBE-compliant and a STROBE/RECORD checklist is available as part of the supplementary material.

### Setting

Connecticut is a high-income state with marked geographic and socioeconomic disparities, including substantial overdose burden in several urban centers and variable prevention infrastructure across urban, suburban, and rural communities. Consequently, it has substantial geographic variation in overdose burden across urban, suburban, and rural communities.

Overdose surveillance and response activities involve CT DPH, local health districts/departments, emergency medical services, law enforcement, harm reduction organizations, and treatment programs. Alerts from the Overdose Mapping Application Program [ODMAP] and other overdose-related communications are used to identify overdose spikes, disseminate situational awareness and coordinate response across these sectors.

### Survey development

In consultation with substance use and harm reduction experts, we developed a brief exploratory survey focused on four domains: communication receipt, perceived utility of communication channels, organizational response actions, and internal receipt of CT DPH overdose alerts. We selected 7 professionals working in New Haven County to pilot the survey and share their feedback. These were individuals whose jobs involved overdose prevention and response and who came from different professional backgrounds. We solicited feedback from these professionals on the phrasing of each question-and-answer choice and used their comments to develop the final survey.

### Recruitment and eligibility

Participants were eligible if they were 18 years or older and held jobs that responded to and/or prevented overdoses in Connecticut. We initiated recruitment through opioid task forces and professional networks and encouraged forwarding to additional relevant stakeholders to broaden representation across sectors and regions. We also held presentations and conversations with task force members in order to encourage them to take the survey.

Because no statewide roster exists for professionals involved in overdose prevention and response, this approach produced a convenience sample rather than a representative statewide sample. Survey participation was voluntary and participants were not compensated for their time. As a result, our study size was dictated by the number of professionals who voluntarily elected to take the survey. The initial recruitment wave in January-February 2024 did not target a specific professional group or county. After reviewing early responses, we conducted additional targeted outreach from July-December 2024 to first responders and respondents from Fairfield County to improve descriptive coverage of these groups. Because recruitment was adaptive and non-probability based, sampling may be biased in a way that cannot be accounted for, and the sample should not be interpreted as representative of all overdose prevention and response professionals in Connecticut.

### Survey content

The survey had seven questions administered through Qualtrics **[Table S1**] and is available to view at the following link: https://yalesurvey.ca1.qualtrics.com/jfe/form/SV_1TT2xyabKh2wPiK. We captured basic demographic information, specifically where respondents worked, how their organization was funded, and what counties their organization served. We focused on state funding because it could directly impact receipt of overdose-related communications from the state. We asked participants to report which forms of information they received about opioid-involved overdoses in their community, which form they perceived as most helpful, how their organization responds to overdoses or overdose clusters, and who in their organization receives overdose alerts from CT DPH. We chose these questions specifically because there are many ways by which professionals may learn about overdoses and there is no comprehensive listing of these different forms of communication, and out of those forms it is unclear what people actually find useful.

There is no comprehensive listing of the ways in which organizations respond to overdoses and we therefore sought also to better understand this. Finally, there was also some concern in New Haven that ODMAP spike alerts from CT DPH were only reaching leadership and not necessarily being shared with client-facing staff.

### Data aggregation

We analyzed question responses using descriptive statistical measures in Qualtrics Microsoft Excel and R. We recoded responses into aggregated categories and stratified survey items by these categories for analysis. Specifically, we aggregated jobs, forms of communication and forms of overdose response **[Tables S2-S4]**.

We conducted the aggregation and stratification in Microsoft Excel for simplicity and then transferred the resulting tables to R for visualization. Aggregated categories were used to improve interpretability of visualizations and provide a larger sample size for stratified analyses. When possible, we recoded free-text ‘other’ responses into predefined job categories using an a priori coding scheme, then stratified responses by these categories for analysis **[Table S5]**. We specifically recoded respondents’ professions when possible. Most “other” responses lacked enough detail to be reliably recoded.

### Preference-Receipt Mismatch Index

We created a novel exploratory index, that we termed the Preference-Receipt Mismatch Index [PRMI], as a heuristic measure of the gap between the percent of choices identifying an aggregated communication category as most helpful and the percent of choices reporting receipt of that category **[Formula 1]**.

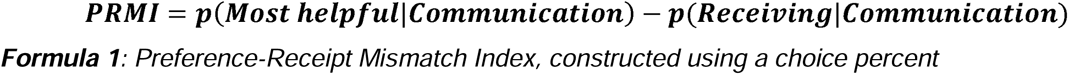

The PRMI was used descriptively and was not intended as a validated scale or inferential measure. Positive values indicate that the percentage of choices identifying a communication category as most helpful exceeded the percentage of choices reporting receipt of that category. Negative values indicate that reported receipt exceeded perceived utility. Because PRMI is exploratory and has not been validated, we interpret it only as a descriptive indicator of possible mismatch, not as an inferential measure or measure of communication effectiveness.

### Choice of Qualtrics metric

Qualtrics offers a choice percent and response percent for multi-select questions. For multi-select questions, we use choice percent, defined as the number of times an option was selected divided by the total number of selections made for the question. This is necessary since we use aggregated categories and this metric ensures that we do not double count since the percentages add up to 100% regardless of aggregation. For demographic information, we use response percent, defined as the number of respondents selecting a choice divided by the total number of respondents. While this metric may double count when using aggregated categories and artificially inflate figures as a result, the demographic analysis is unaggregated so there is no concern about double counting. A response percent is also a more interpretable metric when understanding participant characteristics.

### Data and code availability

Deidentified survey data, aggregation tables and code to produce data visualizations are all available at our Github repository: https://github.com/a-ram-1/overdose_communication_utility

## Results

### Participant information

The survey had 156 respondents in total, out of which 149 completed the survey and therefore provided responses that were usable for analysis. Respondents were considered eligible if they were above the age of 18 and had a job that was involved in overdose prevention or response. All counties were represented among the respondents. All Connecticut counties were represented. The largest proportions of respondents reported working in Hartford, New Haven, and Fairfield counties, which are also counties with the highest fatal overdose burden in the state. The sample was not, however, designed to produce county-representative estimates; county-stratified findings should therefore be interpreted descriptively **[Table S6]**.

Counties with higher overdose burden [Fairfield, Hartford, New Haven] were slightly underrepresented [e.g. 22.18% of respondents work in Hartford County, which had 27.95% of all fatal overdoses in 2024], whereas counties with lower overdose burden [Middlesex, Tolland, Litchfield, Windham] were overrepresented [e.g. Middlesex County had 3.66% of all fatal overdoses in 2024, but 10.91% of respondents reported working in Middlesex County]. This distribution provided responses from all counties, but county-level findings should be interpreted descriptively because recruitment was not designed to produce county-representative estimates.

Respondents reported a range of professional roles, allowing us to describe how reported communication receipt, perceived utility, and response actions varied across job categories. Local ambulance/EMS services and harm reduction organizations/SSPs were tied for the most responses [42], with local health department/district and substance use treatment programs being the third and fourth most common choices **[Table S7]**. 25 respondents chose an “other” category, which consisted of entries such as “Prevention”, “Pharmacy” and “RBHAO.” When possible, these entries were aggregated with other job categories.

Finally, respondents reported having multiple sources of funding. Two EMS participants did not provide funding information. We focused on obtaining information about state funding to see if receiving state funding affects an organization’s propensity to receive alerts and other overdose-related information from the state. Respondents most frequently received funding from the CT DPH [40.7%], with all other forms of funding [Connecticut Department of Mental Health and Addiction Services, other harm reduction funding, no harm reduction funding] at roughly equal proportions **[Table S8]**.

### Communication receipt and perceived utility

We first assessed whether professionals reported receiving different overdose-related communication channels and which channels they perceived as most helpful. We compared reported receipt and perceived utility of communication categories, then used PRMI as an exploratory summary of potential mismatch between what respondents received and what they identified as most helpful. We stratified survey results by county, job and funding source and calculated the PRMI in all cases using the aggregated communication categories. We found that responses were most variable when stratified by profession.

Using choice percentages, peer information sharing accounted for the largest share of reported communication receipt. Less than 15% of selected choices indicate receipt of any other form of communication. However, because these are choice percentages from multi-select responses, they should not be interpreted as the percentage of respondents receiving each communication type. Stratifying by profession showed a similar trend with reported receipt – less than 20% of selected choices report receipt of any form of communication **[Figure 2]**.

**Figure 2:**
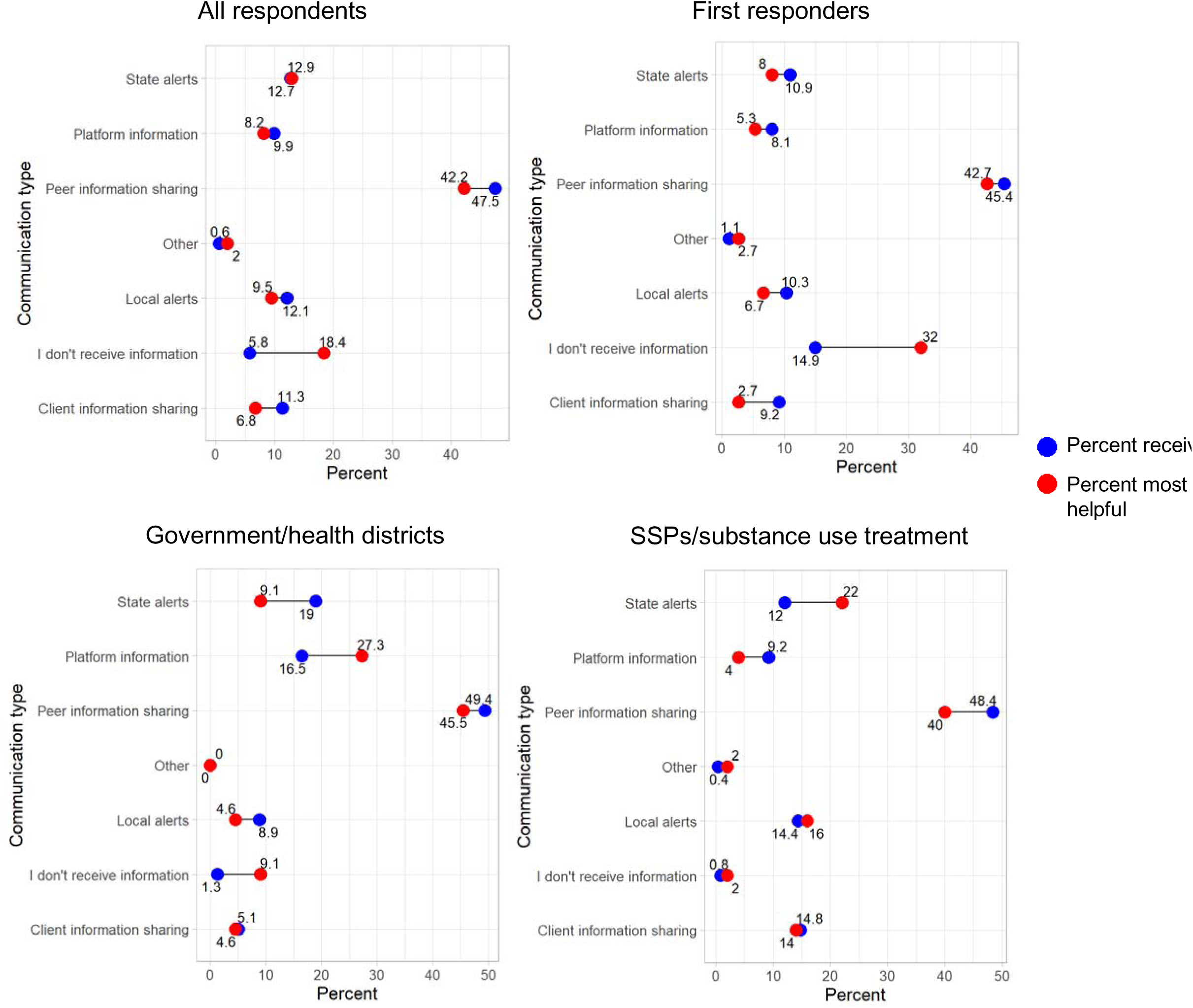
Communication form helpfulness v. receipt [choice percent]

Additionally, we identified profession-specific preferences for communication. We found that SSPs and substance use treatment programs prefer state alerts while finding peer informat on sharing to be less helpful, despite often being the ones to disseminate peer information. On the other hand, government and health districts – a category mostly comprised of respondents from local health departments and health district as opposed to state government--found state alerts to be less helpful, while preferring platform information. They found local alerts, which they often generate, to be more helpful than state alerts but less helpful than platform information. Many claimed that they do not receive overdose information. Among first responders, “we do not receive information” was selected relatively often in the perceived utility item. Because this response does not represent a communication channel, we interpret it cautiously as suggesting either limited receipt of overdose-related communications, low perceived relevance of available channels, or ambiguity in the survey response option.

These results are somewhat driven by the fact that different jobs receive forms of communication at different frequencies, with respondents tending to prefer forms of communication that they receive. First responders revealed that they overwhelmingly do not receive overdose-related information, with 14.9% of choices indicating a lack of receipt.

Government/health district workers received more platform information [16.5% of choices] than other professions. Notably, SSPs reported receiving local alerts more frequently than the government/health district workers whose organizations may be issuing the alerts, suggesting that local communication pathways may be uneven across sectors. Additionally, SSPs/substance use treatment received state alerts less frequently than government/health district workers [12% of choices v. 16.5% of choices].

These findings are recapitulated using the PRMI using responses stratified by profession. Peer information sharing had negative PRMI values across professional groups, indicating that reported receipt exceeded selection as the most helpful category. In contrast, several formal or platform-based communication categories had positive PRMI values within specific professional groups, suggesting possible gaps between channels perceived as useful and channels reported as received **[Figure 3]**.

**Figure 3:**
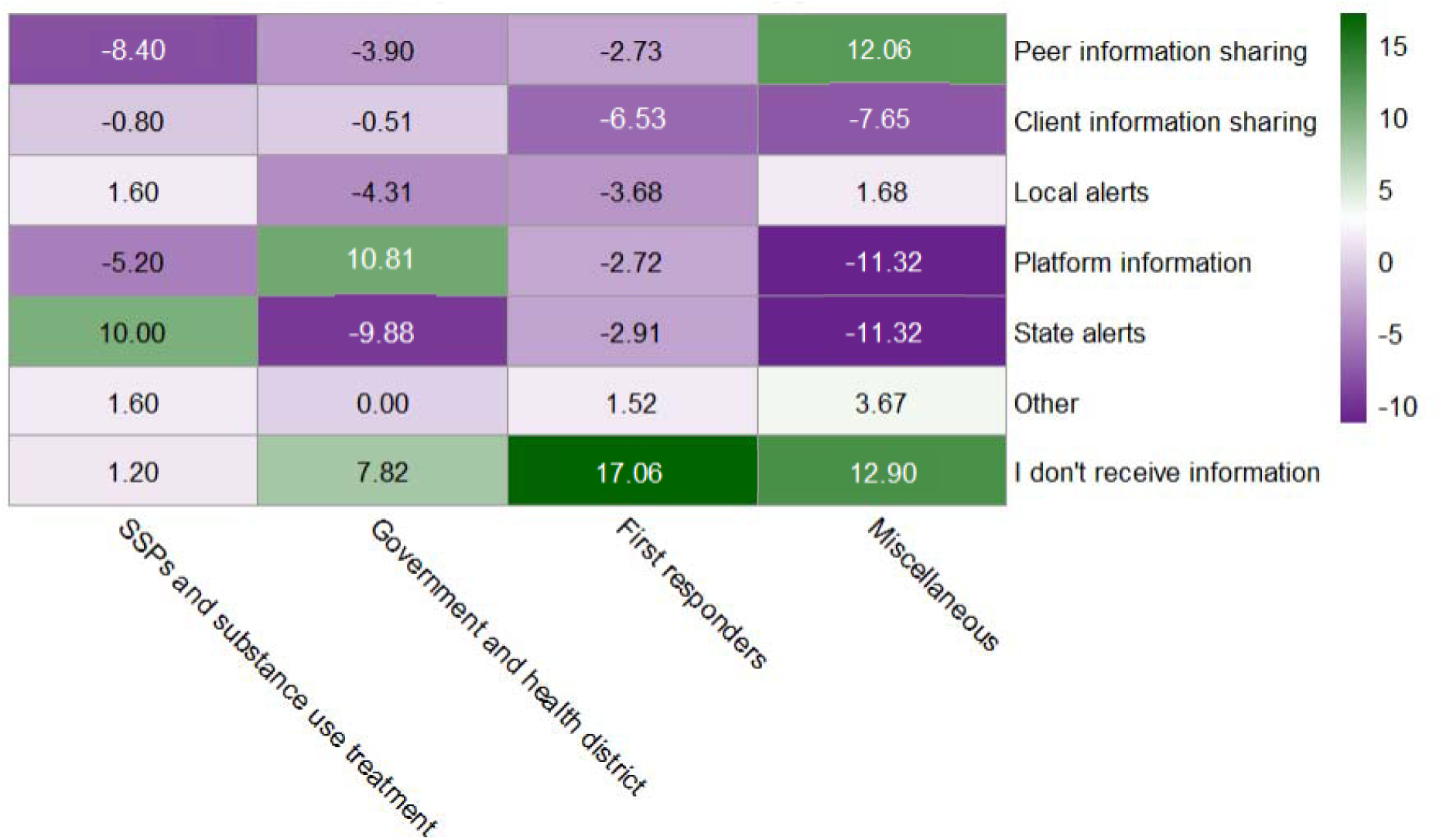
Preference-receipt mismatch index by job [constructed using choice percent]

Stratifying by county and funding source yielded fewer distinctions. Stratifying by county showed that there were few distinctions among larger counties, but smaller counties had more variable PRMI values. “We don’t receive information” had positive or zero values for all counties and client information sharing had negative values for all counties **[Figure S1]**.

Stratifying by funding source revealed that state alerts had the highest PRMI value for DPH-funded organizations, suggesting that DPH-funded organizations were more likely than others to prefer DPH alerts. Additionally, organizations with “other” and “no” funding had higher PRMI values for “we don’t receive alerts” than DPH and DMHAS-funded organizations, suggesting that they are much less likely to receive other information about overdoses **[Figure S2]**.

We obtained additional granularity on each profession’s communication preferences by identifying the most commonly received form of communication for respondents preferring a given form of communication **[Figure 4]**.

**Figure 4:**
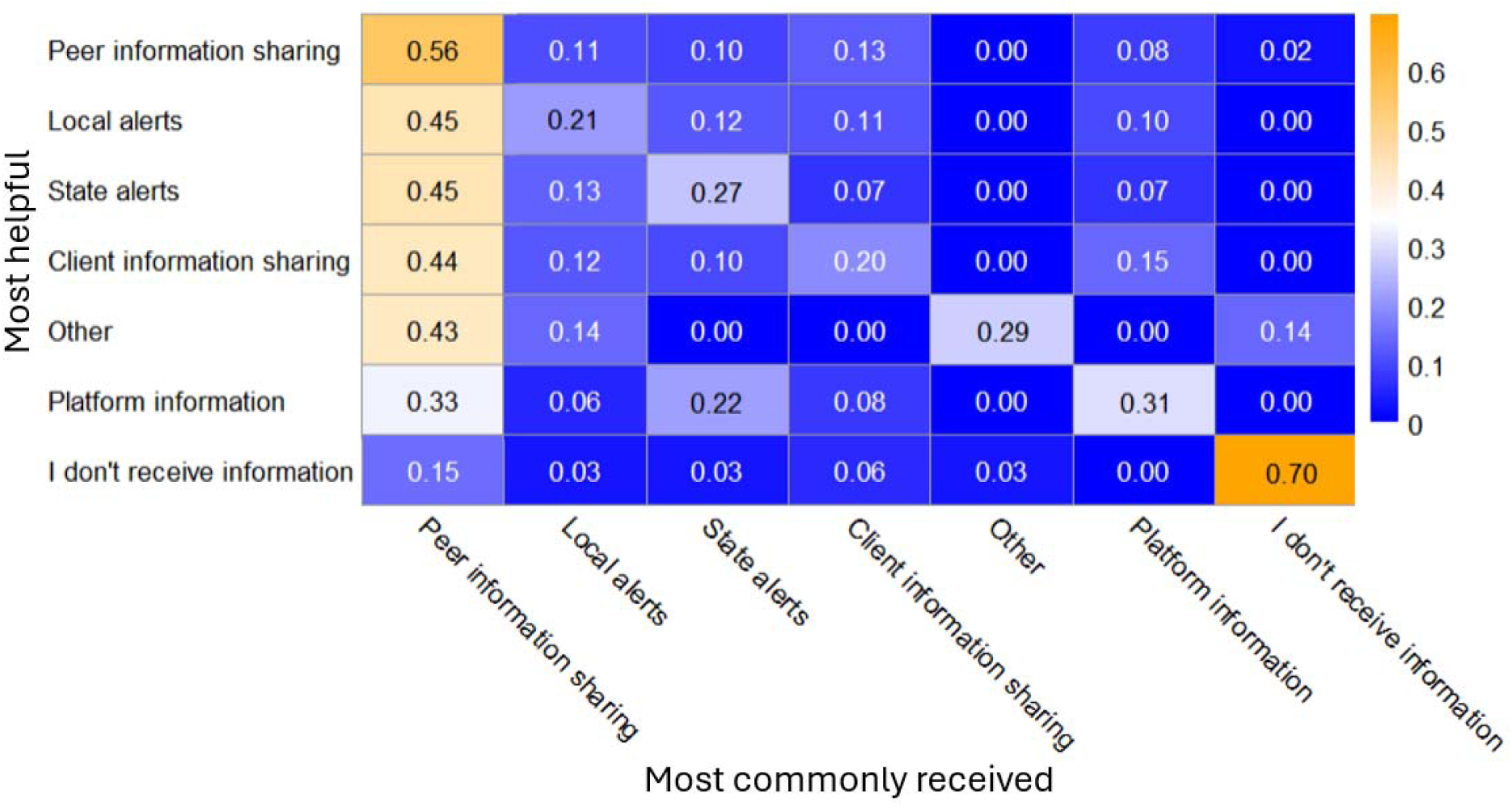
Communication form receipt stratified by most helpful form of communication [choice percent]

Stratifying in this manner suggests several patterns. First, respondents preferring peer information sharing appear to receive other forms of communication at a low frequency compared to others. As a result, peer information sharing likely provides them with most of their overdose-related information. Second, respondents preferring information from clients also appear to receive other forms of communication, aside from peer information sharing, at a low frequency compared to others. Third, when there is another form of communication that respondents receive aside from peer information sharing and communication from clients, they appear to prefer it. Respondents who prefer local alerts, state alerts and platform information appear to do so despite the fact that they receive peer information at a much higher frequency. Across communication categories, 28 respondents selected as most helpful a form of overdose-related communication that they did not report receiving. This finding supports the interpretation that some professionals may value communication channels that are not consistently reaching them.

### Responses to overdoses and overdose clusters

Because professional groups play different roles during overdose events, we also described reported organizational responses by professional role. We investigated the different ways that organizations could respond to overdoses using aggregated response categories. We specifically evaluated clinical and operational changes [expecting more patients, elevating response protocols], informing the public, outreach to people who use drugs [PWUD] and surveillance/investigation [investigating reasons for overdoses, increasing surveillance of affected areas]. All aggregated job categories reported multiple forms of overdose response, although the distribution of response types differed by professional role **[Figure 5]**.

**Figure 5:**
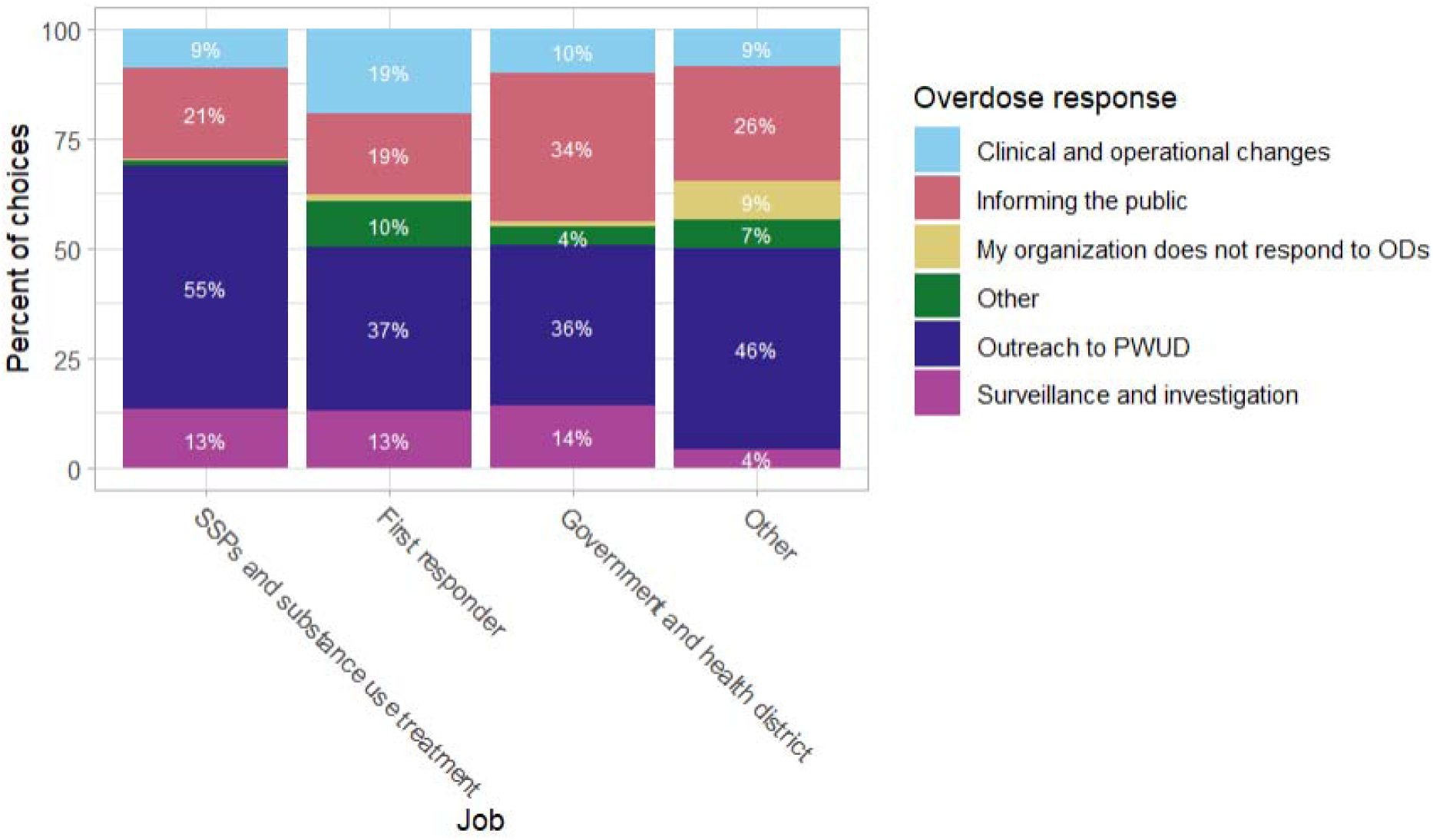
Overdose response by job category [choice percent]. “Other” represents heterogeneous roles and is presented for completeness without inference.

SSPs and substance use treatment and first responders mostly perform outreach to PWUD [55% of choices], government/health districts perform roughly equal amounts of outreach to PWUD and the public [36% and 34% of choices respectively], and other jobs that could not be categorized into these classes mostly reach out to PWUD [46% of choices]. SSPs perform the most outreach to PWUD. All professions that we were able to categorize perform roughly equal amount of surveillance. First responders made the most operational changes [19% of choices] and did the least amount of outreach to the public [19% of choices]. Some respondents from all professions did report that they did not respond to overdoses [less than 1% of choices for all professions besides “other”, which had 9% of choices], but SSP/substance use treatment program workers were the least likely to do this [0.37% of choices].

These results further demonstrate a distinction between professions – in addition to having different preferences for overdose communication and receiving different forms of communication at different frequencies, they also engage in various forms of overdose response at different frequencies.

### Within-organization dissemination of alerts

Limited reported receipt may reflect not only external dissemination gaps, but also internal dissemination processes within organizations. We used CT DPH’s ODMAP spike alerts as a case study to evaluate this. Unlike other forms of communication, these alerts have a standardized format and are sent out as emails to a variety of organizations statewide.

Because CT DPH ODMAP spike alerts are a standardized state communication channel, we used them as a case example to examine reported internal dissemination within organizations. This analysis should be interpreted descriptively because the survey did not directly measure alert list membership, timing, forwarding practices, or whether recipients acted on specific alerts. We find that alerts reach a wide variety of staff within an organization and that client-facing staff among SSPs and first responders report alert receipt more often than leadership **[Figure 6]**. Notably, among first responders we found that 21.4% of choices indicate that respondents are not sure who received these alerts within their organization.

**Figure 6:**
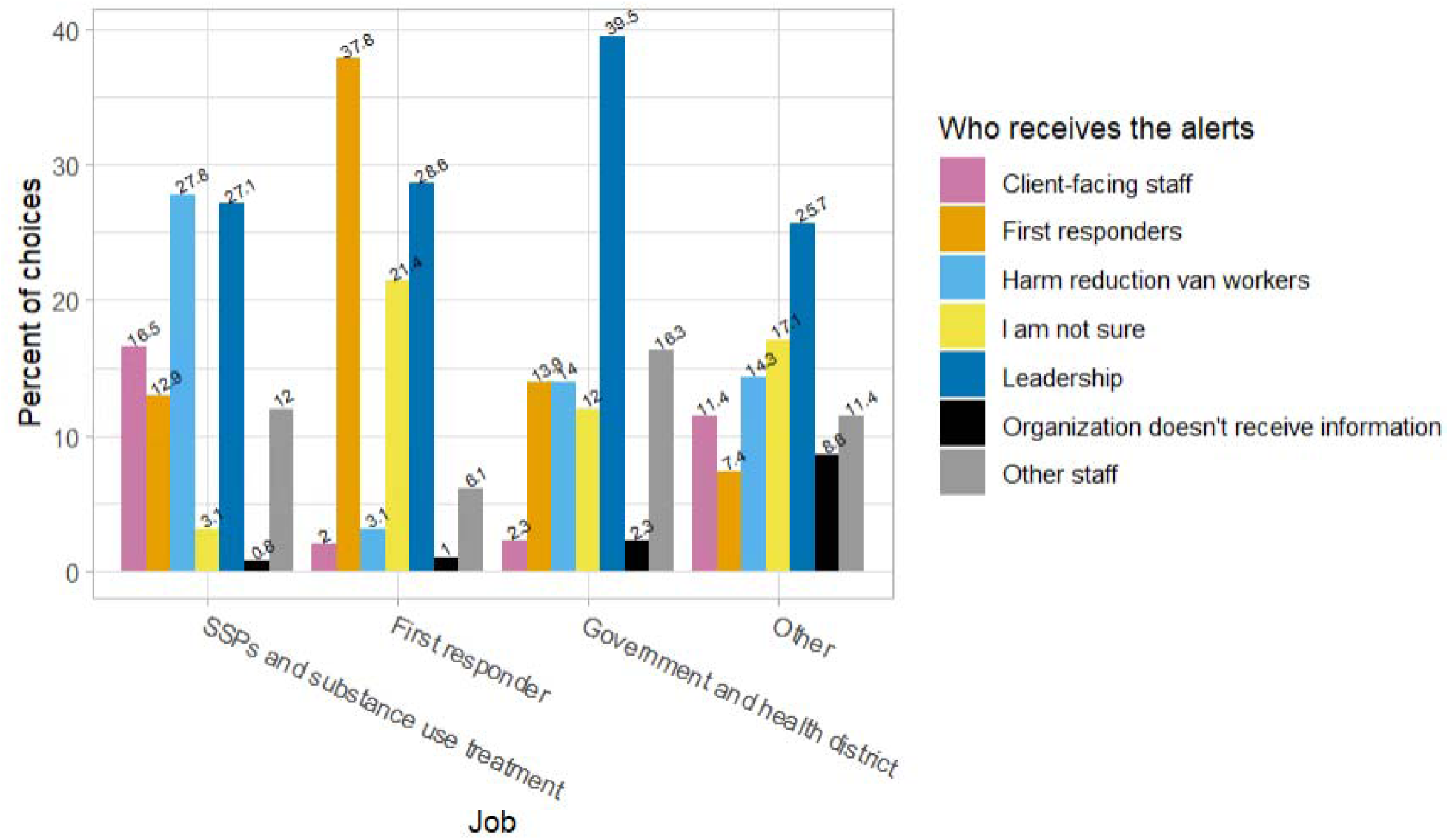
CT DPH ODMAP spike alert receipt by job category [choice percent]. “First responders” comprises the categories “first responders”, “medical providers”, “paramedics”, “emergency department” and “emergency dispatch”.

## Discussion

This exploratory survey suggests that Connecticut’s overdose communication ecosystem is role-dependent and unevenly distributed across professionals involved in overdose prevention and response. Communication systems may support overdose response when timely, trusted, and role-relevant information reaches professionals positioned to act. My findings suggest that informal peer networks are widely used, formal alert receipt is uneven, and reported response actions differ by professional role. We uncovered a landscape in which information preferences vary by profession and a major way professionals respond to overdose-related communication is by reaching out to people who use drugs. We also identify that this information landscape is disjointed, with respondents reporting limited receipt of all forms of communication except peer information sharing. These insufficiencies in reach appear to be addressed through widespread peer communication networks, which may not necessarily be seen as helpful compared to other forms of information, but have the advantage of being far more readily available.

We interpret these findings through a practical public health communication lens focused on four descriptive domains: reported reach of overdose-related information, perceived utility of communication channels, internal dissemination within organizations, and reported response actions. We do not interpret these survey findings as evidence that any communication channel caused specific organizational responses or overdose outcomes.

### Communication receipt and perceived utility differ by professional role

One of the main results we observed was that profession, as opposed to county or funding sources, was the main differentiating factor when it came to what sort of information respondents received, what they found helpful and how they responded to overdoses. We interpret “most helpful” as a measure of perceived utility, not as a direct measure of adoption or effectiveness.

My analysis does not directly measure adoption or effectiveness of communication channels. Instead, the “most helpful” item should be interpreted as perceived utility among the communication options listed. This distinction is important because respondents may receive and use multiple channels even if they identify only one as most helpful.

Of note is the finding that the forms of communication that an organization most readily adopts [the forms of communication that they find most helpful], are often not the most common forms of communication they receive. This can lead to an information landscape where professionals may receive more information than they find useful. In settings where staff already face high workload, poorly targeted or low-actionability communications may contribute to alert fatigue; however, this study did not directly measure alert burden, timeliness, or response delay.

Overall, this points to the need to not overwhelm professionals with overdose-related communication, but rather to tailor communication to people’s needs. This necessarily involves understanding what needs professionals have and developing or tailoring existing communication approaches to better suit them. Role-tailored opt-in communication pathways may help balance reach with relevance by allowing professionals to receive communications that match their responsibilities, preferred format, and desired frequency. Because this study did not directly measure alert burden or alert fatigue, this recommendation should be tested in future work.

The data also paints a portrait of a disconnect between first responders and other professions in that EMS do not necessarily receive overdose-related information, and different communication forms reach first responders far less efficiently than other professions. This may, however, not be an issue requiring a solution. When we attended task force meetings to recruit survey participants, EMS mentioned that OD communication and alerts are not timely enough to be useful for their day-to-day activities, and that leadership does not necessarily share information about overdoses unless it relates to an emerging crisis or trend of concern. This pattern should not be interpreted as negligence or exclusion. First responders may need different types of overdose information than public health, harm reduction, or treatment professionals. For example, real-time emergency dispatch and field experience may be more relevant for immediate response, whereas trend alerts may be more useful for planning, outreach, staffing, or public communication. Overdose information about trends and other alerts is often not more useful to EMS than the information they receive simply from a day on the job.

### Formal state and local communication channels may not consistently reach all intended audiences

Using choice percentages, communication categories other than peer information sharing accounted for smaller shares of reported receipt choices. This pattern suggests that formal communication channels may have uneven reach, although the magnitude should be interpreted cautiously because choice percentages are not respondent-level prevalence estimates. There are many possible causes of this issue. When we were recruiting for the survey, we conducted informal interviews with task force participants, as well as EMS at the CME course at which we presented. They brought up several points that may aid in interpretation of these results -- first, some were confused about how to receive CT DPH spike alerts because they found the website confusing. It is unclear how many people find these websites confusing but this in itself could pose a barrier to reach. Platform information from data sources such as ODMAP is also often highly proprietary and many organizations do not necessarily qualify to view this data even if they could benefit from it.

This inaccessibility can limit the alerts’ efficacy and impede people who may benefit from these tools from accessing them. The more available a tool is, the more easily it can reach populations that can benefit from it. The Litchfield County Opiate Task Force, for example, has information about overdose spikes on their website as well as an opt-in text alert system accessible to the public.^11^ This initiative provides widespread access to valuable information produced by ODMAP, which is a highly proprietary tool designed to be inaccessible to anyone who is not part of an approved list of agencies that has specifically signed an agreement with HIDTA, a federal anti-drug trafficking program.^12^ Improving accessibility of these tools is one way to help improve their reach.

### Informal networks provide valuable information

A look into the different forms of communication using the Proctor’s Implementation Outcome construct *appropriateness* sheds light on the value of informal communication networks created through peer information sharing.^10^ The findings are consistent with a communication ecosystem in which informal peer and client/community networks help compensate for gaps in formal alert reach, timeliness, trust, or local specificity. If forms of communication aside from peer information sharing are available, respondents tend to prefer it. Combined with the fact that 28 respondents also report preferring a form of communication that they do not appear to receive, we can see that existing gaps in communication systems are currently being filled through peer-led means but would perhaps be more effectively addressed by improving reach of more formal communication channels such as state/local alerts.

While the data also reveals that participants may prefer forms of information other than peer and client information sharing, it is crucial to not understate the importance of these forms of information. Especially given the stigmatized nature of drug use, official channels [e.g. state/local health department] are not always the most appropriate ways to gather information about emerging crises. People who use drugs often hesitate to call 911 because they worry that EMS work with law enforcement and may be undocumented, have active warrants or have other reasons to want to avoid law enforcement. People who use drugs may hesitate to go to drug testing sites out of a fear of prosecution, but might be willing to tell harm reduction van workers about a strong batch they heard has been causing several overdoses, or that they are experiencing unwanted effects when using fentanyl adulterated with certain other substances. This is valuable information that may otherwise be inaccessible. Aside from information from people who use drugs themselves, peer information is also highly capable of providing on-the-ground, hyper-local details about emerging crises that can easily be translated into action.

Additionally, sometimes the infrastructure simply does not exist for there to be, for example, overdose alerts from the local health department. Several respondents reported not receiving ODMAP spike alerts, which may reflect differences in enrollment, list maintenance, role turnover, or dissemination practices. These operational factors may contribute to gaps in reach even when communication tools exist. Particularly when facing these structural barriers, peer/client information is a necessity. State alerts, while available statewide, also do not necessarily offer the sort of hyper-local geographic specificity that communities may need for overdose prevention/response.

### Organizations may lack clear internal dissemination pathways

Using Proctor’s Implementation Outcomes as a guideline, our analysis reveals some issues with information *penetration*. One of the reasons we may be seeing limited reported information receipt is because these different forms of overdose-related communication may be unable to penetrate fully into some organizations. This signals that these organizations may lack standardized ways to disseminate information from leadership to frontline staff. Using CT DPH ODMAP spike alerts as an example, we found that harm reduction organizations and EMS reported a roughly equal proportion of leadership and frontline staff receiving CT DPH alerts, while government/health districts reported that mostly leadership receive DPH alerts.

While this suggests that CT DPH alerts are able to penetrate into organizations and reach a wide variety of staff for EMS and SSPs/substance use treatment, the same may not be the case for other forms of communication. Further analysis is required to better understand the penetration of other forms of overdose-related communication, but a lack of penetration to client-facing or frontline staff can impact efficacy of overdose response/prevention activities. It is striking that some respondents report not necessarily receiving updates and information about overdoses despite having professions that involve overdose prevention/response. In some cases, such as with first responders, it is not always useful for leadership to share more updates about overdoses with client-facing staff, but depending on an organization’s needs this may be helpful.

### Limitations

Due to the small sample size and large number of response choices per question, we were not able to run any factor analysis on our survey and lack statistical power to make more quantitative claims. Because the study was exploratory, used a convenience sample, and included multiple response options with small subgroup counts, we did not conduct inferential comparisons. Due to our use of a convenience sample, our findings may be biased but we are unable to identify the potential direction or magnitude of any bias. Findings should be interpreted as descriptive patterns that can guide hypothesis generation, system improvement, and future qualitative or mixed-methods work. We can say that the generalizability of our results, however, is limited by the fact that we had a small sample size that was also a convenience sample.

Another limitation of our analysis is our choice of metric. Using a choice percentage means that the proportions we see in our figures are artificially deflated -- as a result reported receipt of different forms of communication is still limited but likely not as bad as the numbers themselves may make it seem. If we disaggregate the communication categories and look at response percentage, we can see that an issue still persists, but is less severe than the analysis would make it appear to be **[Figure S3]**.

The choice percentage also makes PRMI more challenging to interpret, but the PRMI should be used more as an exploratory indicator of a discrepancy as opposed to an exact measure. We decided that, despite its issues, a choice percentage is still useful. While it has a slightly more challenging interpretation than response percentage, it was preferable to a measure that will double-count respondents and overinflate figures, in addition to making interpretation challenging in its own way by creating aggregate proportions that add up to more than 1.

### Future directions

This analysis lays the groundwork for further investigation into design and improvements to existing overdose communication systems, including but not limited to spike alerts, overdose surveillance software such as ODMAP or EpiCenter and more informal approaches such as task force emails and updates from local SSPs and drug checking sites. Future research should emphasize user-centered design and qualitative and mixed-methods approaches to examine why different professional groups prefer different communication channels, what makes overdose alerts actionable, how information moves within organizations, how much alert fatigue professionals experience and how people who use drugs can be meaningfully included in overdose communication systems.

Additionally, the communication systems we evaluate are all intended for professional use, but there is no reason why people who use drugs should not have access to these same resources, especially given that they are the ones who are at risk of overdose and death. Incorporating people who use drugs more meaningfully into the overdose communication landscape and providing them with easier access to overdose-related communication tools, however, requires working with them to understand their needs and preferences. The more robust an overdose communication system is, and the better it meaningfully reaches all relevant members of a community, the more effectively everyone can help respond to a crisis and work to prevent the next one. This is why co-designing these systems with end users in mind is crucial, since they need to be tailored to their preferences.^92,93^ The more an end user is incorporated into the creation and maintenance of an alert system, the more likely they are to perceive it as useful.

These findings suggest four practical improvements that could be made to overdose communication infrastructure in Connecticut. First, we propose developing opt-in communication and alert systems that are tailored to different professions, both in terms of frequency of communication and platform through which the information is communicated [e.g. email versus text or phone call]. This helps ensure that the information that professionals receive is genuinely helpful to them, is accessible on their preferred communication platform and does not result in alert fatigue.

Second, we recommend ensuring that mailing lists and other communication systems are easily accessible, since professionals sometimes struggle to access information like CT DPH ODMAP spike alerts. It may even be useful to have a directory online with a list of opt-in communication systems listed by location so professionals can easily find resources that relate to their communities.

Third, in order to address the discrepancy between leadership and other staff receiving DPH spike alerts, we suggest implementing standardized dissemination approaches tailored to different professions that either instruct leadership on disseminating information to their staff or ensure that DPH is able to directly reach staff as needed. Not every staff member benefits from receiving spike alerts, but it is important to make sure that alert penetration is not unnecessarily limited within an organization and that staff who could benefit from receiving alerts are in fact able to receive them.

Finally, we also advocate for the use of standardized alert-to-action playbooks. All professions surveyed are performing several different activities when responding to overdoses, and it is unclear how they prioritize these different actions or if they adhere to standardized protocols when responding to an emergent crisis. Creating standardized protocols for overdose response helps ensure timely action when it comes to the care they provide their patients and clients, since there is no need to decide how to prioritize different response activities. In all cases, we strongly suggest that any proposed improvements to protocols and communication systems be made using extensive feedback and close collaboration with the target population, if it is not possible for the target audience to make these improvements themselves.

## Abbreviations

SSP: syringe service program
CT DPH: Connecticut Department of Public Health
ODMAP: Overdose Detection Mapping Application Program
OD: overdose
EMS: emergency medical services
PWUD: people who use drugs
PRMI: Preference-Receipt Mismatch Index

## Supporting information

STROBE/RECORD checklist

