## Supplemental tables and figures for "Overdose-related communication in Connecticut: reported receipt, perceived utility, and responses among overdose prevention and response professionals"

| **Question** | **Type of answer choice** |
| --- | --- |
| Where do you work? | Choose multiple |
| What counties does your organization serve? | Choose multiple |
| Please check all of the following methods by which you receive updates and information about opioid-involved overdoses and overdose clusters in your community. | Choose multiple |
| Which of the following in this list have you found to be MOST HELPFUL in terms of responding to opioid-involved overdoses in your community? | Choose one [response options are the same as the previous question] |
| How do you/your organization respond to opioid-involved overdoses in your county? | Choose multiple |
| Who in your organization receives updates and information about opioid-involved overdoses and overdose clusters in your community from the Connecticut Department of Public Health [CT DPH]? | Choose multiple |
| Please check the funding sources your organization receives for your overdose prevention and response efforts. | Choose multiple |

**Table S1**: All survey questions

| **Job category** | **Job** |
| --- | --- |
| First responder | Fire department |
|  | Police department |
|  | Local ambulance/EMS |
|  | Hospital |
| Government and health districts | Local government |
|  | Local health department/district |
|  | State/federal government |
| SSPs and substance use treatment | Harm reduction organization/SSP |
|  | Substance use treatment program |
| Other | Other [write-in category] |

**Table S2**: Job category aggregation scheme

| **Communication category** | **Form of communication** |
| --- | --- |
| Local alerts | ODMAP spike alert emails from local health governance [e.g. local health department/health district] |
|  | ODMAP spike alert text messages from local health governance |
|  | Phone calls from local health governance |
| Peer information sharing | Regular messages [e.g. listserv] from local first responder agency |
|  | Regular messages [e.g. listserv] from opioid task force, prevention councils or other source |
|  | Text messages about overdoses [not from ODMAP] |
|  | Phone calls about overdoses [not from ODMAP] |
|  | Communication with colleagues |
|  | Insights from harm reduction services |
|  | Insights from nearby drug testing sites |
| State alerts | ODMAP spike alert emails from Connecticut Department of Public Health [CT DPH] |
|  | Phone calls from CT DPH |
| Client information sharing | Communication with patients/clients |
| Platform information | Review of ODMAP or other overdose data [e.g. EpiCenter] |
| we don’t receive updates or information about overdoses | we don’t receive updates or information about overdoses |
| Other | Other [write-in category] |

**Table S3**: Communication category aggregation scheme

| **Overdose response category** | **Type of overdose response** |
| --- | --- |
| Outreach to people who use drugs [PWUD] | Increased harm reduction supply distribution in affected areas [naloxone, fentanyl test strips etc] |
|  | Notify people who use drugs about recent overdoses |
|  | Provide people who use drugs with harm reduction education ["don't use alone and go slow"] |
|  | Refer patients/clients to local drug testing sites |
| Informing the public | Inform the public about substances causing overdoses [e.g. fentanyl-contaminated crack] if known |
|  | Inform the public about overdoses |
| Clinical and operational changes | Be on higher alert for an influx of patients |
|  | Modify or elevate preemptive overdose prevention protocols |
| Surveillance and investigation | Increased surveillance of affected areas |
|  | Investigate reasons for recent overdoses |
| My organization does not respond to overdoses | My organization does not respond to overdoses |
| Other | Other [write-in category] |

**Table S4**: Overdose response category aggregation scheme

| **Free text "other" response** | **Recoded value** | **Rationale for recoding** |
| --- | --- | --- |
| American Medical Response Waterbury | First responder | American Medical Response is a first responder agency |
| Volunteer fire dept | First responder | Fire departments are first responders |
| Hospital based EMS service | First responder | EMS services are first responders |

**Table S5**: “Other” response recoding

| **County** | **Percentage of choices** | **Fatal overdoses per 10000 residents** | **Percentage of 2024 fatal overdoses** |
| --- | --- | --- | --- |
| Hartford | 22.18% | 3.05 | 28% |
| New Haven | 20% | 3.16 | 27.2% |
| Fairfield | 13.8% | 1.62 | 16.2% |
| New London | 12.4% | 3.65 | 10% |
| Middlesex | 10.9% | 2.21 | 3.7% |
| Litchfield | 8.7% | 1.86 | 3.5% |
| Tolland | 6.2% | 1.55 | 2.4% |
| Windham | 5.8% | 2.37 | 2.9% |

**Table S6**: County-based distribution of responses v. 2024 fatal overdose distribution [choice percent]

| **Job** | **Number of respondents** | **Percentage of respondents** |
| --- | --- | --- |
| Local ambulance/EMS service | 42 | 28% |
| Harm reduction organization/SSP | 42 | 28% |
| Local health department/district | 18 | 12% |
| Substance use treatment program | 15 | 10% |
| Fire department | 11 | 7.3% |
| Hospital | 11 | 7.3% |
| Police department | 4 | 2.7% |
| State/federal government | 4 | 2.7% |
| Local government | 3 | 2% |
| Other | 25 | 16.7% |

**Table S7**: Respondents by profession [response percent]

| **Funding source** | **Number of respondents** | **Percentage of respondents** |
| --- | --- | --- |
| Connecticut Department of Public Health [CT DPH] | 54 | 38.60% |
| Connecticut Department of Mental Health and Addiction Services [CT DMHAS] | 45 | 32.14% |
| Other harm reduction funding | 42 | 30% |
| My organization does not receive external funding for overdose prevention | 48 | 34.29% |

**Table S8**: Funding received by respondents [response percent]. Two participants declined to respond.

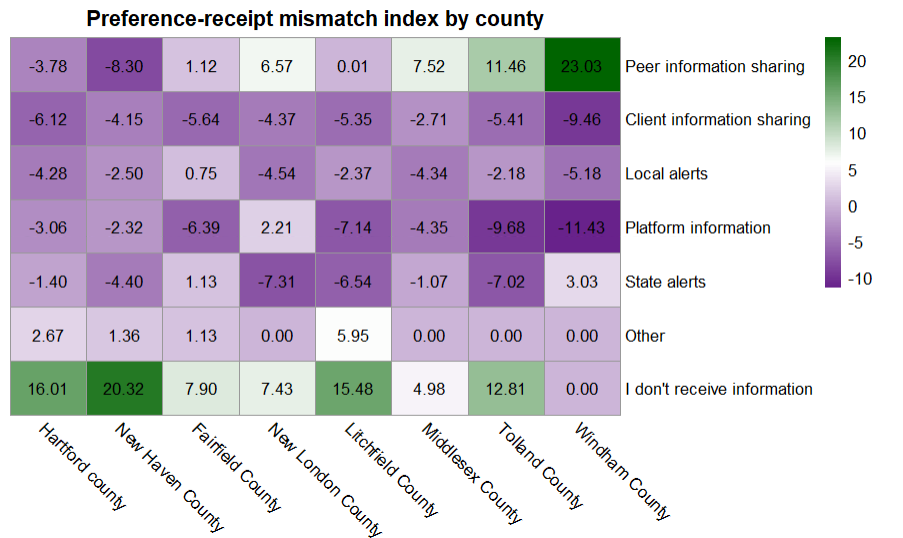

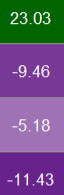

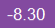

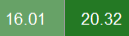

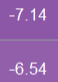

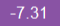

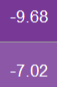

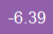

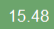

**Figure S1**: Preference-receipt mismatch index by county [constructed using choice percent]

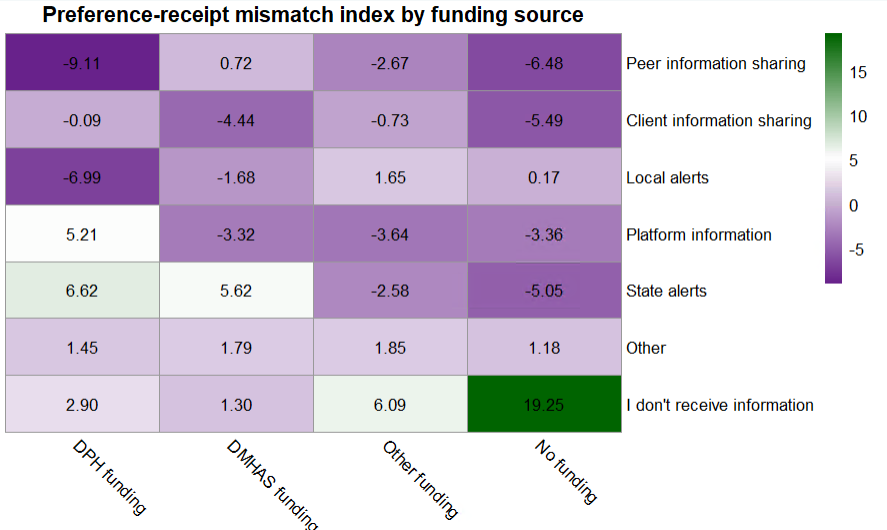

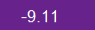

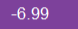

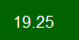

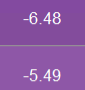

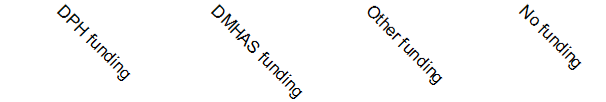

**Figure S2**: Preference-receipt mismatch index by funding source [constructed using choice percent]

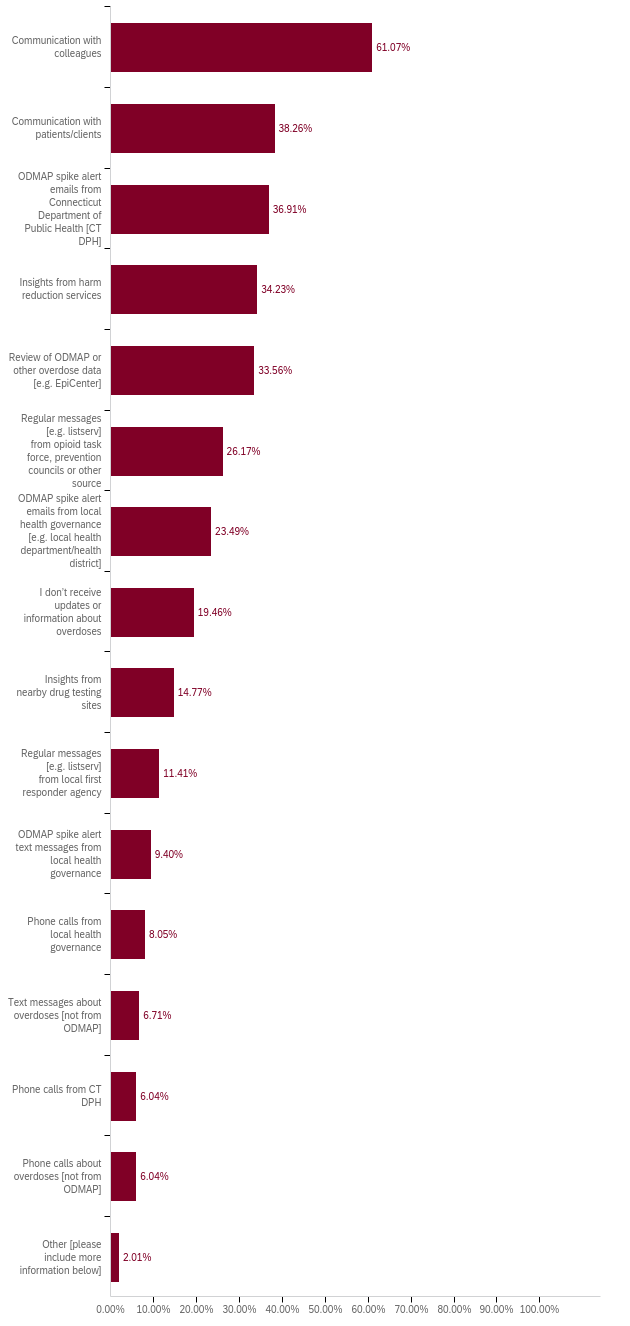

**Figure S3**: Percent of respondents that received different forms of overdose-related communication [response percent]
